# Real-world brain imaging in a population-based cohort enables accurate markers for dementia

**DOI:** 10.1101/2025.02.13.25322207

**Authors:** Reijo Sund, Juho Seppänen, Elaheh Moradi, Sami Väänänen, Jani Miettinen, Juhana Hakumäki, Toni Rikkonen, Heikki Kröger, Heli Koivumaa-Honkanen, Alina Solomon, Jussi Tohka, Alzheimer’s Disease Neuroimaging Initiative

## Abstract

**INTRODUCTION:** While a vast amount of MRI data are collected for healthcare delivery, generating real-world evidence (RWE) in Alzheimer’s and related diseases (ADRD) research is substantially limited by lack of methods and results showing how routine MRIs can be used for ADRD imaging studies.

**METHODS:** We compared three established ADRD biomarkers (total gray matter, hippocampal and ventricular volumes) in four groups (normal, subjective complaints, mild cognitive impairment, and dementia) between the general population of women born in 1932-1941 in the Kuopio region of eastern Finland (population-based OSTPRE cohort, N=14220) and a well-characterized research cohort (ADNI).

**RESULTS:** A total of 2434 brain MRIs for 1885 women were collected between 2003-2022 by the public healthcare provider covering all residents in the region. The established biomarkers were overall aligned between these cohorts.

**DISCUSSION:** Typical biomarkers extracted from real-world brain MRI scans collected over 20 years are suitable for generating RWE in ADRD research.

**Highlights:**

- Real-world brain MRI is applicable for generating evidence in ADRD research
- First study comparing a real-world MRI cohort with an established research cohort reference
- A methodological framework for RWE ADRD studies using routinely collected MRIs

## 1 Background

Magnetic resonance imaging (MRI) plays a crucial role in the diagnostic and prognostic evaluation of neurodegenerative diseases. Following recent regulatory approval of amyloid-targeted disease-modifying therapies for Alzheimer’s disease (AD), MRI has also become important for assessing eligibility for treatment and monitoring the effects including adverse events [1]. In non-pharmacological interventions aiming to reduce the risk of dementia, MRI can provide very useful information about potential intervention effects on the brain and the window of opportunity for prevention [2,3]. However, the number of MRIs in clinical trials and research cohorts is limited. This limitation poses a significant challenge for developing predictive biomarkers using artificial intelligence (AI)-based methods. Furthermore, research cohorts often exhibit inherent selection biases and lack the necessary population heterogeneity [4–7].

In contrast, a vast amount of real-world imaging data (RWD) is collected routinely for delivery of health care. For instance, in the United Kingdom alone, over 700,000 head MRIs are performed annually [8]. Similarly, elderly people in the United States undergo head MRI at a rate exceeding 40 scans per 1000 persons annually [9]. Surprisingly, the utilization of RWD in dementia research remains an underexplored area. While some studies have focused on memory clinic populations (e.g. [10–13]), there exists a wealth of untapped potential in mining the picture archiving and communications systems (PACS) at hospitals [14].

One of the key limitations for generating RWE in Alzheimer’s and related diseases (ADRD) research is the absence of methods, protocols and evidence for the potential to use clinical MRIs for dementia imaging studies. The inherent difficulties associated with clinical MRIs—such as non-standardized pulse sequences, varied acquisition protocols, suboptimal data quality, and potential artifacts— require evidence supporting their value as imaging biomarkers. This challenge is not unique to MRIs; similar obstacles persist in the broader context of medical RWD utilization [15].

In this study, we compare ADRD biomarkers derived from routine brain MRI scans extracted from the PACS of a large regional public healthcare provider in Eastern Finland with a well-established reference research cohort, Alzheimer’s Disease Neuroimaging Initiative (ADNI), where imaging has followed a strict protocol. We also describe the steps needed to locate and organize RWD for MRI scans, link them to diagnostic data from medical records, and compute and quality-check the relevant biomarkers to ensure their suitability for generating RWE in ADRD research.

## 2 Methods

### 2.1 The OSTPRE population-based cohort and real-world MRI scans

The Kuopio Osteoporosis Risk Factor and Prevention Study (OSTPRE) is a population-based cohort including all women born in 1932-1941 who lived in the Kuopio County in Eastern Finland in 1989 (N=14 220). The use of total population guarantees the diversity, equity and inclusion of the target population. These women have been followed up every fifth year since 1989 using questionnaires (and measurement visits for a subpopulation). Several randomized controlled trials (RCTs) and other studies with additional data collection have also been conducted within the cohort. The OSTPRE study originally aimed to investigate factors associated with bone mineral density, bone loss, falls and fractures, but the scope has broadened to healthy aging in general. National and local register data have been linked to the data set, including e.g. health care admissions since 1969 and clinical radiologic imaging data since 2003. The OSTPRE cohort has been previously described in detail [16–19]. The study has been approved by the Ethics Committee of Kuopio University Hospital. Permissions for register data have been obtained from the Finnish data permit authority for the social and health care data (Dnro THL/6840/14.02.00/2020). The study is performed in accordance with the ethical standards as laid down in the 1964 Declaration of Helsinki and its later amendments.

MRIs are routinely stored to a regional PACS on the Wellbeing services county of North Savo, Kuopio, a large regional public healthcare provider in Eastern Finland. PACS started to serve Kuopio University Hospital in 2003, and district hospitals and primary care units joined the regional PACS between 2006-2011. Between 2003-2023, the regional PACS included 294 045 MRI examinations, of which 100 379 covered the head/brain region. Of the head/brain MRI examinations, 52 780 were performed for patients aged over 50 years, and the yearly number tripled after the first years (Supplementary Figure 1). Of these examinations, 44 513 were conducted in the radiology department of Kuopio University Hospital, 3 448 in privately operated mobile MRI units visiting district hospitals in the neighboring towns of Iisalmi and Varkaus, and 4 819 in stationary privately operated MRI units (Supplementary Table 1). Most of the MRI scans were ordered by neurology clinics (Supplementary Table 2). The MRIs for the OSTPRE cohort were obtained from the regional PACS.

### 2.2 Extraction of the real-world brain MRIs

A flowchart of the routine brain MRI scans for OSTPRE cohort participants for the years 2003-2022 is shown in Figure 1. All head area MRI scans of the OSTPRE participants were exported from the regional PACS in which the imaging scans were stored as DICOMs (Supplementary Text 1). First, PACS reporting database was used to create a list of examinations of the OSTPRE participants. The list was fed to a custom script which tagged the examinations in the PACS database. The images of the tagged examinations were then sent from PACS using teleradiology to a server running storescp application from the DICOM Toolkit (version 3.6.7) [20]. A python script run by PACS before teleradiology sent pseudonymized images by erasing DICOM tags including patient identification information (e.g. name and personal identification code assigned to each Finnish resident). The server with storescp running stored the images to a network attached storage (NAS) device. Data were organized so that all images belonging to the same examination visit were stored in their own subfolder. These subfolders were compressed with 7-zip and then moved to the secure research environment of the OSTPRE study. Basic metadata about the examinations were extracted from reporting databases of PACS and Radiology Information System (RIS) and from the DICOM tags of exported image files.

**Figure 1.**
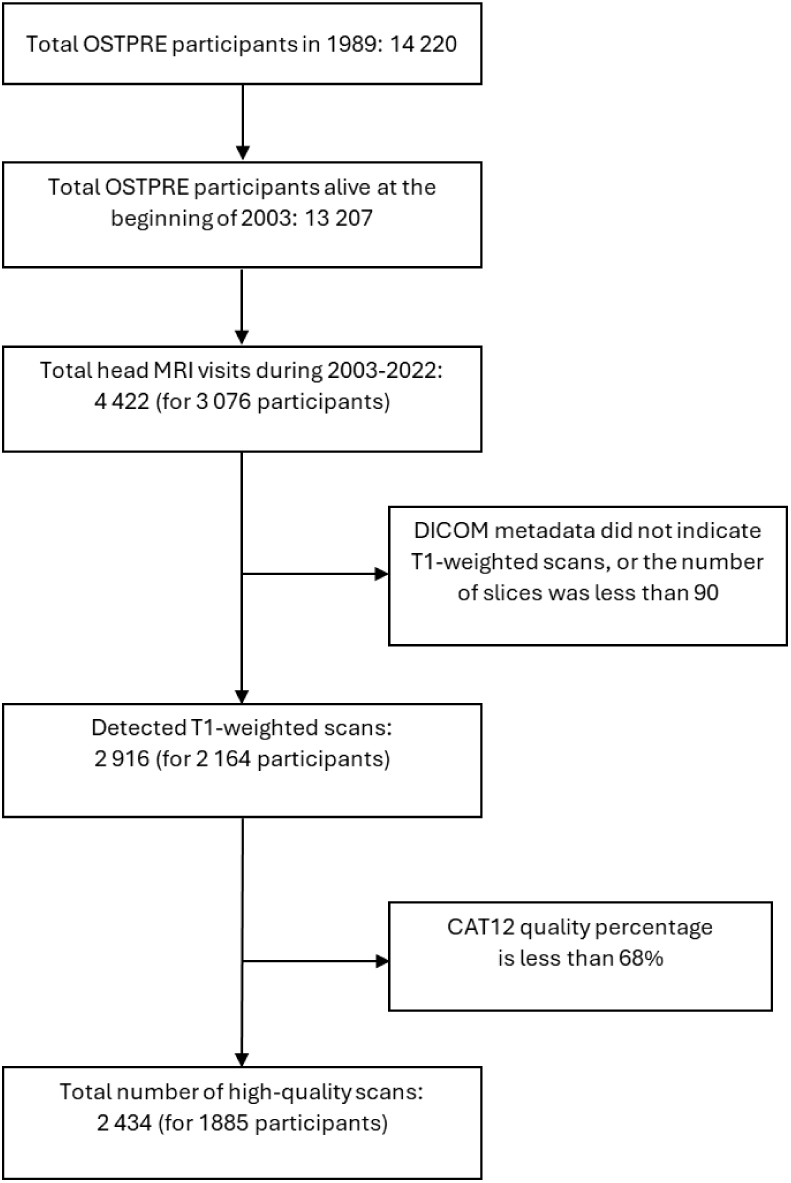
Flowchart of the real-world brain MRI scans for OSTPRE cohort participants

Basic metadata were used to identify MRI examinations of the head area using Finnish radiologic procedure codes (AA1BG, AA1CG, AA1DG) with a structure similar to the NOMESCO Classification of Surgical Procedures (NCSP) [21,22]. These examinations were decompressed from the 7-zip archives and reorganized so that each OSTPRE participant with head area MRI scans had a folder with subfolders for each separate examination visit, containing the actual 2.3 million DICOM-files (separate files, not directories; in scans each slice is one file). Next, DICOMs were converted to Neuroimaging Informatics Technology Initiative (NIfTI)-format and stored in a folder structure similar to DICOMs by using dcm2niiX version v1.0.20220720 [23,24]. The number of converted NIfTI files corresponding to separate scans was 59 039. Information about the scanner manufacturer and the used field strength was extracted from the metadata tags.

T1-weighted scans were selected from NIfTI files and stored them according to Brain Imaging Data Structure (BIDS, see supplementary text 1) standard simplifying the image analyses [25,26]. No simple way to automatically identify the MRI sequence type exists for the real-world clinical data acquired using various protocols and scanners [27]. Therefore, we developed an algorithm that first determined if the image was acquired using T1-weighting (T1) without contrast agent by applying regex search (see supplementary Text 1; for inclusion we used “mpr|mprage|t1_se|sag|tra|T1|TFE” and for exclusion “gd”) to the DICOM metadata tag “ProtocolName”. Additionally, it was checked that the scan had sufficiently high resolution and coverage of the brain region based on the number of slices (at least 90). After the images were organized into the BIDS, a comprehensive validation script was executed to ensure the integrity and consistency of the data format [28].

### 2.3 Processing of MRIs

The T1-weighted MRIs were segmented into grey matter (GM) and white matter (WM) and non-linearly normalised by using default preprocessing of the CAT12 Toolbox [29], Version 8.1, under Matlab 2022b and SPM12 [30]. The preprocessing utilised the unified segmentation [31] to remove B0-inhomogeneities and produce an initial segmentation that was used for (local) intensity scaling and adaptive non-local means denoising [32]. An adaptive maximum a posteriori [33] segmentation with a hidden Markov random field [34] and partial volume effect model [35] was used for the final segmentation. For the non-linear registration to the MNI152Nlin2009cAsym template, the Shooting method [36] with modulation was used. Finally, neuromorphometrics atlas was used to define the ROI volumes [37].

CAT12 yields automatic quality control measures ([29], supplement 4) and the image quality percentage of 68% was considered to be sufficient for the measures to enter the analysis. We additionally performed the MRI volumetry by another pipeline, SynthSeg [38], version 2.0, and used a robust regression technique to identify the measures that were not concordant and therefore needed to be removed from the analysis. Briefly, a robust correlation using bisquare weighting function was performed using MATLAB (robustfit function) and outliers were defined as those images with residuals larger than 8 times median absolute deviation estimate of the standard deviation of the residuals (Supplementary Figure 3).

We considered total gray matter, hippocampus (left and right) and ventricle (left and right lateral and inferior lateral ventricles) volumes as MRI biomarkers, because they have clear established links to dementia/AD, and have been previously used as secondary endpoints in AD-related randomized controlled intervention trials (e.g., [2,39]).

### 2.4 Identification of cognitive status from real-world health data

To determine the cognitive status of the OSTPRE participants at the time of each brain MRI visit, all records potentially reflecting cognitive issues were first identified from the Hospital Discharge Register (1969-1993), the Care Registers for Health and Social Welfare Care (1994-), the Causes of Death Statistics (1972-), Register of Primary Health Care Visits (2011-), Special Reimbursement Register (1964-) and Drug Purchases (1993-) using relevant codes (Supplementary table 3). Cognitive status was categorized as no memory complaints (NMC) subjective memory complaints (SMC), mild cognitive impairment (MCI) or dementia.

For each MRI visit, the cognitive status was determined based on the visit date versus register code dates. A participant was categorized as having SMC, MCI or dementia if at least one relevant diagnostic code was recorded in at least one register before the MRI visit date. If none of the relevant codes were recorded before the MRI visit date, the participant was categorized as NMC. If a milder degree of cognitive impairment at the time of the MRI visit was followed by more pronounced impairment within one year after the MRI visit, the participant was categorized as having more pronounced impairment at the MRI visit, i.e. SMC followed by MCI within one year was categorized as MCI and MCI followed by dementia within one year was categorized as dementia.

### 2.5 The ADNI research cohort as reference for the OSTPRE real-world MRIs

The ADNI (The Alzheimer’s Disease Neuroimaging Initiative) was launched in 2003 as a public-private partnership, led by Principal Investigator Michael W. Weiner, MD. The primary goal has been to test whether serial MRI, positron emission tomography (PET), other biological markers, and clinical and neuropsychological assessment can be combined to measure the progression of MCI and early AD. For up-to-date information, see (www.adni-info.org) [40].

ADNI participants were selected to match the OSTPRE cohort by sex, ethnicity, and cognitive status categories. As ADNI participants were typically younger than OSTPRE participants, we did not match for age, and treated age as a covariate in the statistical analyses.

For this study, only MRI scans acquired at the ADNI baseline visit were considered. We used the same MRI processing pipeline for ADNI as described above for OSTPRE. Cognitive status at the baseline visit was determined by the ADNI Clinical Core based on cognitive assessments, mainly the Mini-Mental State Examination (MMSE) and Clinical Dementia Rating (CDR). Supplementary table 3 describes the ADNI cognitive status categories.

### 2.6 Statistical methods

The associations between the volume measure of interest (total gray matter, hippocampus, ventricles) and cognitive status in OSTPRE and ADNI data were analyzed using a linear regression model. The standardized volume measure was used as a dependent variable in the model and categories of cognitive status and cohort were the main independent variables. Also age, manufacturer, field strength as well as total intracranial volume (TIV) were included as independent variables to adjust for variation in background factors. All brain volume measures were standardized to have mean zero and standard deviation one to achieve comparable effect sizes. Potential non-linearity for age and TIV was accounted for by using cubic B-splines for those terms in the regression model. As there were several MRIs for some OSTPRE participants, sandwich estimators were used to adjust for clustering by participant. Predictive margins of measures for cognitive status and cohort were used in the comparisons of contrasts of interest (between cognitive status categories within and between OSTPRE and ADNI). Statistical modeling was conducted in R v4.3.2 [41] using package ggeffects v1.5.2 [42] for determining predictive margins and adjusted contrasts. Package dplyr v1.1.4 [43] was used for data preparation and package ggplot2 v3.5.1 [44] for visualization.

## 3 Results

Population characteristics for the two cohorts are shown in Table 1 and Supplementary Figure 2. For the included 1885 women from the OSTPRE cohort (2434 T1-weighted MRI scans) there were 916 comparable women in the ADNI cohort (916 T1-weighted MRI scans).

**Table 1.** Population characteristics.

|  | **OSTPRE** | **ADNI** |
| --- | --- | --- |
| **Number of T1-weighted MRI scans (participants)** | 2434 (1885) | 916 (916) |
| **Age, years, mean (SD)** | 77.9 (SD 4.5) | 72.2 (SD 7.2) |
| **Scanner Field Strength** |  |  |
| 3 Tesla | 426 (17.5%) | 647 (70.6%) |
| 1.5 Tesla | 2008 (82.5%) | 269 (29.4%) |
| **Scanner Manufacturer** |  |  |
| Siemens | 1972 (81.0%) | 513 (56.0%) |
| Philips | 442 (18.2%) | 153 (16.7%) |
| GE | 0 (0%) | 250 (27.3%) |
| Toshiba | 20 (0.8 %) | 0 (0%) |
| **Cognitive status** |  |  |
| No memory complaints | 967 (39.7%) | 231 (25.2%) |
| SMC | 479 (19.7%) | 158 (17.2%) |
| MCI | 202 (8.3%) | 380 (41.5%) |
| Dementia | 786 (32.3%) | 147 (16.0%) |

ADNI = Alzheimer's Disease Neuroimaging Initiative, OSTPRE = The Kuopio Osteoporosis Risk Factor and Prevention Study, MCI = Mild Cognitive Impairment, SMC = Subjective Memory Complaints, SD = standard deviation

Figure 2 shows the distribution of raw values for total gray matter, hippocampus, and ventricles volume measures in the OSTPRE and ADNI cohorts. The overall pattern across cognitive status categories for these MRI measures was similar between the two cohorts. However, the OSTPRE cohort exhibited a larger variance.

**Figure 2:**
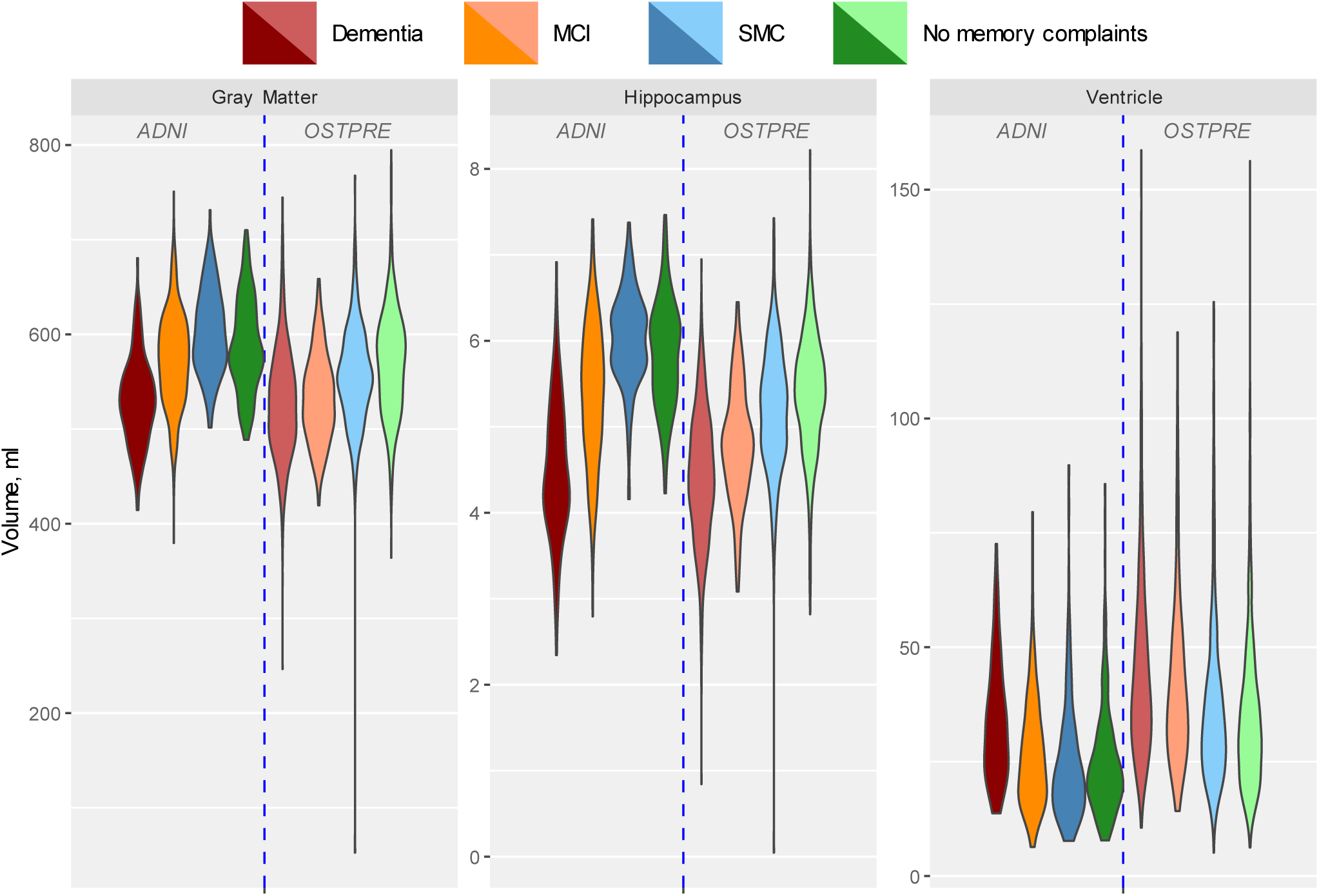
Violin plot of raw values of total gray matter, hippocampus and ventricles volumes across participants, grouped by four cognitive status categories and cohorts (ADNI and OSTPRE). ADNI = Alzheimer’s Disease Neuroimaging Initiative, OSTPRE = The Kuopio Osteoporosis Risk Factor and Prevention Study, MCI = Mild Cognitive Impairment, SMC = Subjective Memory Complaints

In the ADNI dataset, the standardized volume measures were not significantly different between the NMC and SMC cognitive status groups (Table 2, Figure 3). The NMC group was significantly different from both MCI and dementia (P ≤ .002), and MCI was different from dementia (P <.001). In the OSTPRE dataset, NMC and SMC were different in total gray matter and hippocampus volume (P<.001), but not ventricle volume (P=.161). NMC was different from MCI and dementia for all volume measures (P <.001), while MCI was significantly different from dementia for all volume measures except for total gray matter volume (P=.242). In both ADNI and OSTPRE the observed differences were in the expected direction, i.e. NMC≥SMC>MCI>D for hippocampus and total gray matter volumes, and NMC≤SMC<MCI<D for ventricle volume (Figure 3).

**Figure 3.**
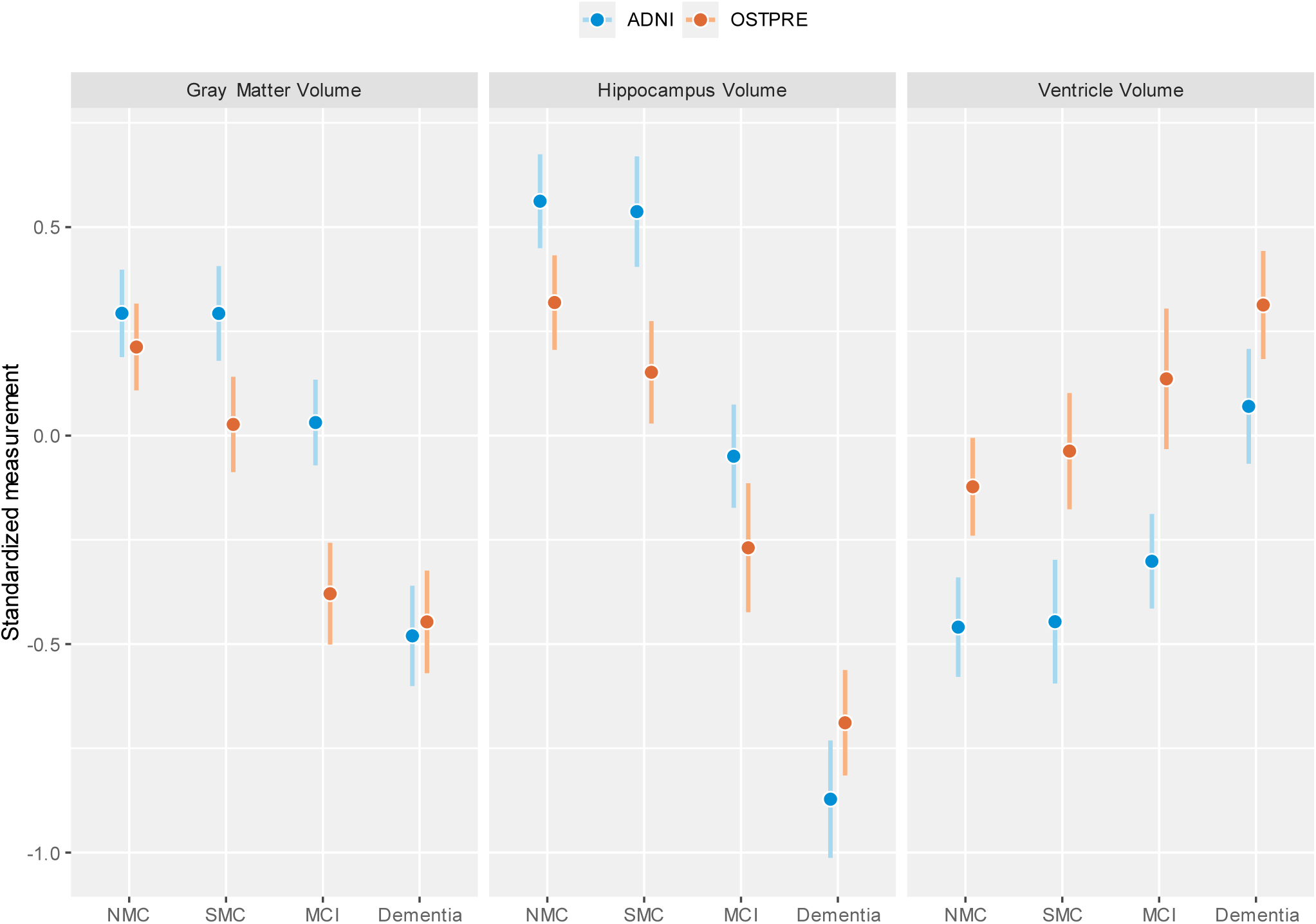
Standardized volume measures with 95% confidence intervals by cohort and cognitive status. NMC=No memory Complaints, SMC=Subjective Memory Complaint, MCI=Mild Cognitive Impairment. ADNI and OSTPRE participants included in analyses are matched by ethnicity, sex, and cognitive status categories. All models are adjusted for age, standardized total intracranial volume (TIV), manufacturer and field strength. ADNI = Alzheimer’s Disease Neuroimaging Initiative, OSTPRE = The Kuopio Osteoporosis Risk Factor and Prevention Study, MCI = Mild Cognitive Impairment, SMC = Subjective Memory Complaints, and NMC = No Memory Complaints

**Table 2.** Standardized volume measures by cohort and cognitive status categories.

|  | ADNI |  | OSTPRE |  | ADNI - OSTPRE differences |  |
| --- | --- | --- | --- | --- | --- | --- |
|  | Between-group difference (95% CI) | P value | Between-group difference (95% CI) | P value | Between-cohorts difference (95% CI) | P value |
| **Total gray matter volume** |  |  |  |  |  |  |
| NMC–SMC | 0.00 (-0.10 to 0.10) | 0.994 | 0.19 (0.10 to 0.28) | < .001 | -0.19 (-0.32 to -0.05) | 0.006 |
| NMC–MCI | 0.26 (0.17 to 0.35) | < .001 | 0.59 (0.49 to 0.69) | < .001 | -0.33 (-0.47 to -0.19) | < .001 |
| NMC–D | 0.77 (0.65 to 0.89) | < .001 | 0.66 (0.56 to 0.76) | < .001 | 0.11 (-0.04 to 0.27) | 0.141 |
| MCI–D | 0.51 (0.40 to 0.63) | < .001 | 0.07 (-0.05 to 0.18) | 0.242 | 0.44 (0.28 to 0.60) | < .001 |
| **Hippocampus volume** |  |  |  |  |  |  |
| NMC–SMC | 0.02 (-0.08 to 0.13) | 0.629 | 0.17 (0.08 to 0.26) | < .001 | -0.14 (-0.28 to -0.01) | 0.040 |
| NMC–MCI | 0.61 (0.51 to 0.71) | < .001 | 0.59 (0.46 to 0.72) | < .001 | 0.02 (-0.14 to 0.19) | 0.779 |
| NMC–D | 1.43 (1.29 to 1.58) | < .001 | 1.01 (0.92 to 1.10) | < .001 | 0.43 (0.26 to 0.59) | < .001 |
| MCI–D | 0.82 (0.67 to 0.97) | < .001 | 0.42 (0.28 to 0.56) | < .001 | 0.40 (0.20 to 0.60) | < .001 |
| **Ventricle volume** |  |  |  |  |  |  |
| NMC–SMC | -0.01 (-0.14 to 0.11) | 0.835 | -0.09 (-0.21 to 0.03) | 0.161 | 0.07 (-0.10 to 0.25) | 0.412 |
| NMC–MCI | -0.16 (-0.26 to -0.06) | 0.002 | -0.26 (-0.41 to -0.11) | < .001 | 0.10 (-0.08 to 0.28) | 0.273 |
| NMC–D | -0.53 (-0.66 to -0.40) | < .001 | -0.44 (-0.54 to -0.33) | < .001 | -0.09 (-0.26 to 0.07) | 0.264 |
| MCI–D | -0.37 (-0.49 to -0.25) | < .001 | -0.18 (-0.33 to -0.02) | 0.026 | -0.19 (-0.39 to 0.00) | 0.053 |

Overall, similarities and differences in MRI volumetric measures between cognitive status groups were comparable between the ADNI and OSTPRE cohorts (Table 2, Figure 3). NMC-SMC differences in total gray matter and hippocampus volumes were more pronounced in OSTPRE compared with ADNI (.19 vs. .00, p=.006 and .17 vs. .02, p=.040). The NMC-MCI difference in total gray matter volume was also more pronounced in OSTPRE compared with ADNI (.59 vs. .26, p<.001). The NMC-dementia difference in hippocampus volume was somewhat more pronounced in ADNI compared with OSTPRE (1.43 vs. 1.01, p<.001). MCI-dementia differences in all volume measures were also more pronounced in ADNI compared with OSTPRE.

Most brain volumetry studies continue to apply linear adjustment for confounding variables (age, sex, TIV) albeit nonlinear relations between confounding variables and brain volumes have been reliably demonstrated [45,46]. We addressed the question in the case of real-world MRI. Figure 4 shows the nonlinear trajectories of MRI measures by age and standardized TIV in a combined cohort of ADNI and OSTPRE. Hippocampus and total gray matter volumes decreased with increasing age, but the slope of decline was less pronounced in the oldest old (80-85+) participants. Similarly, ventricles volume increased with age, with a less pronounced slope of decline in the oldest old. While all MRI volumes increased with increasing TIV, this increase was not linear for hippocampus and ventricle volumes. The TIV increase was more pronounced than hippocampus volume increase, and less pronounced than ventricle volume increase.

**Figure 4.**
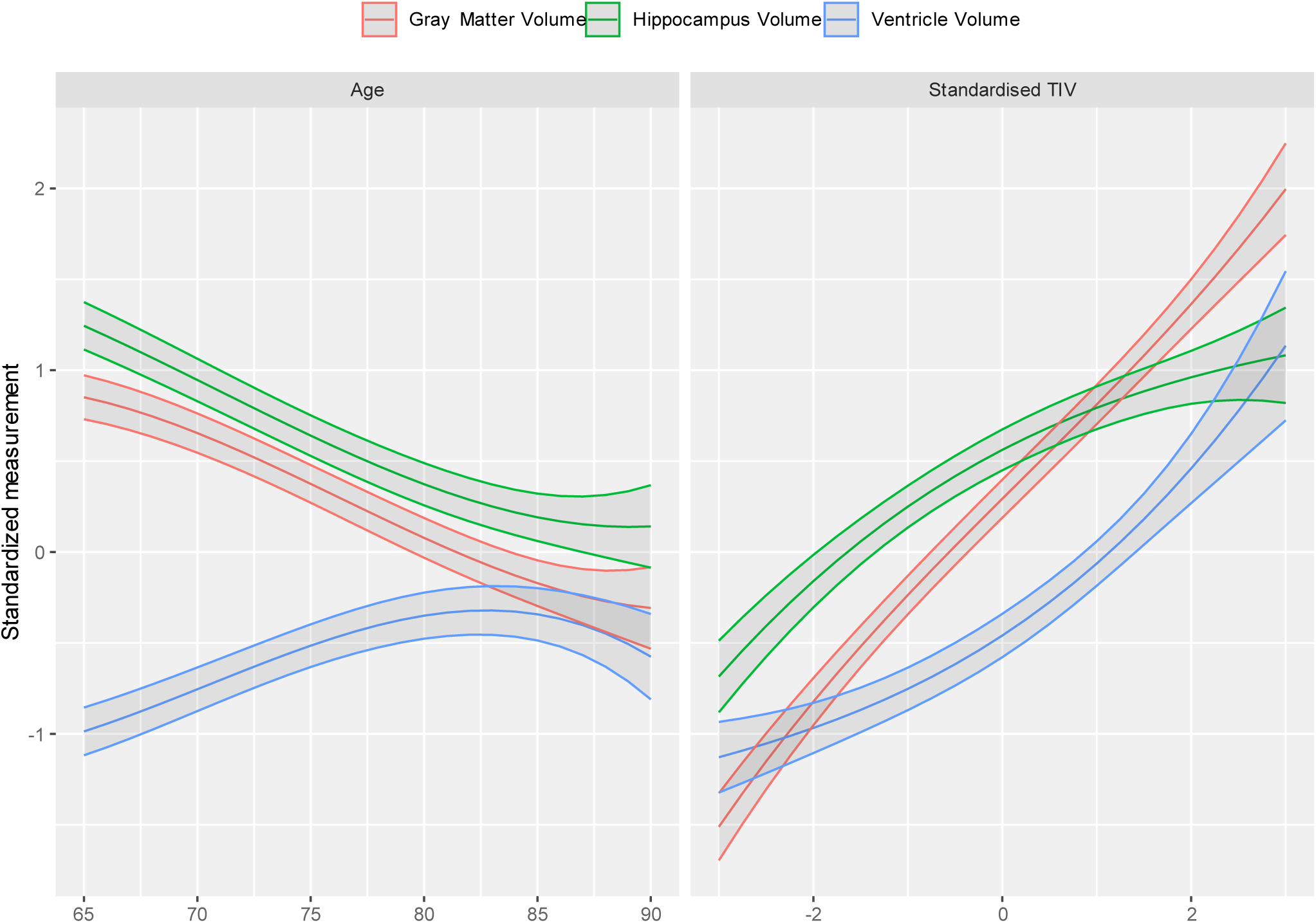
Nonlinear trajectories of MRI measures with 95% confidence intervals by age and standardized TIV in the combined cohort of ADNI and OSTPRE. TIV = Total Intracranial Volume, ADNI = Alzheimer’s Disease Neuroimaging Initiative, OSTPRE = The Kuopio Osteoporosis Risk Factor and Prevention Study

## 4 Discussion

To our knowledge, this is one of the first studies to assess the potential of extracting quantitative biomarkers for AD/dementia from real-world brain MRI scans collected over 20 years (2003-2022). We compared three established dementia-related volumetric measures between four cognitive status groups (normal, subjective complaints, impairments, and dementia) in a large population-based cohort (OSTPRE) versus a well-characterized research cohort reference (ADNI). We focused on total gray matter volume, hippocampus volume and ventricular volumes, because they have well-described links to AD/dementia, and have been previously used as secondary endpoints in AD-related randomized controlled intervention trials [2,39,47–50]. The OSTPRE cohort represents the general population of women born in 1932-1941 in the Kuopio region of Eastern Finland, and the public healthcare provider collecting the MRI scans covers virtually all residents in the region. Thus, the OSTPRE MRI cohort can be considered to represent a RWD source for MRI scans, where MRI examinations are primarily driven by an individual’s healthcare needs.

While the MRI biomarkers were overall aligned between the real-world and research cohorts, they exhibited significant quantitative variations in their capacity to distinguish between different cognitive status groups. The hippocampus volume separated well all groups in both cohorts, except for NMC-SMC comparison in the ADNI cohort. This is a typical finding in research studies [51]. The significant NMC-SMC difference found in OSTPRE but not ADNI is most likely due to the reliance of OSTPRE SMC definition on diagnostic codes from RWD, i.e. the memory complaints were sufficiently pronounced to require a doctor’s referral for examination or at least received doctor’s attention during the health care visit for some other reason. In contrast, SMC in ADNI relied on complaints inquired during voluntary research participation. Ventricle volume separated nearly all cognitive status groups in both cohorts, except for NMC-SMC. As expected based on previous findings [52], between-group differences were smaller for ventricle compared with hippocampus volume, indicating hippocampus volume as a more sensitive dementia-related marker. Similarly to ventricle volume, total gray matter volume separated nearly all cognitive status groups except for NMC-SMC in both cohorts, and MCI-dementia in OSTPRE. MCI as a diagnostic category is much newer than dementia, i.e. older ICD versions did not include codes for MCI, and ICD-10 (introduced in Finland in 1996) has a relatively non-specific MCI code. Diagnostic criteria for MCI have varied considerably since their first introduction in 1998 [53], and the timing and consistency of MCI code use in clinical practice is not fully clear. These differences may also be due to the stricter controlled ADNI imaging protocol, but they may also reflect the participant selection bias inherent to research cohorts, and the specific selection occurring in cohorts tailored for ADRD research. Case-control approaches are particularly prone to overestimating between-group differences [54]. In contrast, population-based cohorts are more likely to reflect the heterogeneity of both populations and MRI methods found in clinical practice.

The distribution of the cognitive status groups showed a higher percentage of dementia cases in the real-world compared with the research cohort. This is not entirely surprising since the RWD was based on the presence of a clinical indication for brain MRI referral. Dementia diagnoses in Finnish registers were previously shown to have good accuracy [55]. Given the limitations of the register-based MCI definition, the population-based cohort included significantly fewer individuals with MCI. Newer diagnostic manuals such as ICD-11 and DSM-5 place more emphasis on this intermediate stage before dementia onset by including a code for mild neurocognitive disorder with more clearly specified criteria [56,57]. As dementia diseases are increasingly diagnosed at earlier stages, diagnostic criteria evolve with the development of more accessible biomarkers (e.g. blood), and disease-modifying therapies become available, the distributions of cognitive status groups in population-based cohorts may also change over time.

To our knowledge, few other studies have focused on extraction of quantitative MRI volume markers from routine MRIs in the context of ADRD research. Two studies assessed RWD data on MRIs in the context of computer aided diagnosis [58,59]. They extracted T1-weighted MRI scans from data warehouse of 39 French hospitals [59] and Research Patient Data Registry of a Mass General Brigham system [58], respectively. Bottani et al. [59] demonstrated the difficulty of classification of persons with dementia diagnosis from persons without diagnoses suggesting dementia and presence of brain lesions (only 64.1% balanced accuracy), a task that is considered “easy” with research cohorts. Leming [58] reported the area under receiver operating characteristics curve (AUC) of 0.82 to separate AD/MCI from healthy controls using convolutional neural network classifier for data including all MRI scan modalities. While data extraction approach especially in [59] features many similarities with ours, including the extraction of routine brain MRI scans from the “regional PACS”, our focus of deriving and comparing quantitative MRI parameters differs from the direct application of machine learning algorithm to imaging data classification of [58,59].

In the realm of medical research, RWD has emerged as a powerful tool. However, its use in the context of MRI is not without challenges. One of the primary hurdles is the quality of RWD MRI itself. Unlike carefully controlled research settings, clinical environments often produce MRI scans with varying degrees of quality. Factors such as patient movement [60] and equipment differences (e.g., between different manufacturers) can introduce artifacts, bias and distortion [61]. Therefore, to ensure data integrity, researchers must implement (automatic) quality control measures [14,29,62], including scripts to check for image quality and standardized protocols for data acquisition. Moreover, the context in which RWD is generated is crucial to understanding its limitations. The referring physicians always have reasons for ordering an MRI scan (and not a CT scan), and the radiologist who reads the requests may use their own judgement as to the chosen modality and the scanner sequences utilized.

Although these situations are mostly covered by mutually accepted protocols, slight deviations can influence the data’s relevance and applicability. This study has demonstrated that RWD MRI data has approximately similar quality than the data from the largest dementia research program. However, getting the MRI data into use requires multiple steps, which are both legal/contractual (agreements) and technical (availability of tools for mass data export from PACS, pseudonymization, format conversion, computational capacity, and data organization). Therefore, careful planning and utilizing existing standards such as BIDS [26] when they exist is crucial.

In conclusion, while RWD offers immense potential for medical research, its effective utilization requires a nuanced approach. Researchers must navigate methodological challenges, address infrastructural limitations, and carefully consider the context in which RWD is generated. By doing so, they can harness the power of RWD to advance our understanding of diseases and develop more effective treatments.

## Data Availability

All data produced in the present study are available upon reasonable request to the authors.

https://github.com/rsund/OSTPRE-brain-MRI

## Code availability

Scripts used for the data analyses are available at: https://github.com/rsund/OSTPRE-brain-MRI

## Author contributions

Conceptualization: all authors; data acquisition and curation: RS, SV, JT, EM, JS and JM; formal analysis: RS, JS, JT, EM and SV; funding acquisition: RS, HK, AS, JT; investigation: all authors; methodology: all authors; project management and operational oversight: RS, SA, HK, JT; scientific oversight: RS, AS, JT, JH, HK, HKH, TR; writing – original draft: RS, AS, JT, SV and JS; writing – review & editing: all authors.

## Acknowledgments

Part of the analyzes were performed on servers provided by UEF Bioinformatics Center, University of Eastern Finland, Finland. This research has been supported by grants 346934, 358944 (Flagship of Advanced Mathematics for Sensing Imaging and Modeling) from the Research Council of Finland; grant 351849 from the Research Council of Finland under the frame of ERA PerMed (“Pattern-Cog”): grant 6671 from Ane and Signe Gyllenberg’s Foundation; and grant 101132933 from the EU Innovative Health Initiative Joint Undertaking (IHI JU) AD-RIDDLE.

Data collection and sharing for this project was funded in part by the Alzheimer’s Disease Neuroimaging Initiative (ADNI) (National Institutes of Health Grant U01 AG024904) and DOD ADNI (Department of Defense award number W81XWH-12-2-0012). ADNI is funded by the National Institute on Aging, the National Institute of Biomedical Imaging and Bioengineering, and through generous contributions from the following: AbbVie, Alzheimer’s Association; Alzheimer’s Drug Discovery Foundation; Araclon Biotech; BioClinica, Inc.; Biogen; Bristol-Myers Squibb Company; CereSpir, Inc.; Cogstate; Eisai Inc.; Elan Pharmaceuticals, Inc.; Eli Lilly and Company; EuroImmun; F. Hoffmann-La Roche Ltd and its affiliated company Genentech, Inc.; Fujirebio; GE Healthcare; IXICO Ltd.; Janssen Alzheimer Immunotherapy Research \& Development, LLC.; Johnson \& Johnson Pharmaceutical Research \& Development LLC.; Lumosity; Lundbeck; Merck \& Co., Inc.; Meso Scale Diagnostics, LLC.; NeuroRx Research; Neurotrack Technologies; Novartis Pharmaceuticals Corporation; Pfizer Inc.; Piramal Imaging; Servier; Takeda Pharmaceutical Company; and Transition Therapeutics. The Canadian Institutes of Health Research is providing funds to support ADNI clinical sites in Canada. Private sector contributions are facilitated by the Foundation for the National Institutes of Health (www.fnih.org). The grantee organization is the Northern California Institute for Research and Education, and the study is coordinated by the Alzheimer’s Therapeutic Research Institute at the University of Southern California. ADNI data are disseminated by the Laboratory for Neuro Imaging at the University of Southern California.

## Consent statement

Informed consent was obtained from all human participants.

## Supplementary Text 1

**PACS** (Picture Archiving and Communication System) is a medical image archiving and communication system that uses the DICOM standard to store, manage, and distribute medical images. PACS systems are used in hospitals, clinics, and other healthcare settings to store and retrieve medical images for diagnosis, treatment, and research. PACS systems typically include a server, workstations, and archive storage devices.

**DICOM** (Digital Imaging and Communication in Medicine) is a standard for storing, transmitting, and communicating medical images and data. It is an international standard that is used by healthcare providers to exchange medical images and data between different imaging modalities, such as X-ray machines, MRI scanners, and CT scanners. DICOM defines a format for encoding image data, as well as a protocol for exchanging images and associated data between devices and networks.

**NIFTI** (Neuroimaging Informatics Technology Initiative) is a file format for storing and exchanging neuroimaging data. It is widely used in the field of neuroscience to store and share brain images acquired from various modalities, such as MRI, fMRI, and CT scans. NIFTI files contain a standardized representation of the image data, including spatial information, intensity values, and header metadata. This makes it possible to easily share and analyze neuroimaging data across different software platforms and research groups.

**BIDS** (Brain Imaging Data Structure) is a data format and workflow for organizing and sharing neuroimaging data. It is a standardized way to store and organize neuroimaging data, including neuroimaging scans, metadata, and derived datasets. BIDS is based on the principles of open science and reproducibility, and it aims to make neuroimaging data more accessible and interoperable.

**regex** (regular expression) is a sequence of characters that defines a search pattern. It is commonly used for string-searching algorithms, “find” or “find and replace” operations on strings, and for input validation. In regex, the vertical bar | is known as the OR operator that makes it possible to match any one of the patterns separated by it. For example, the regex “mpr|T1” will match if either “mpr” or “T1” is found.

**Supplementary Figure 1:**
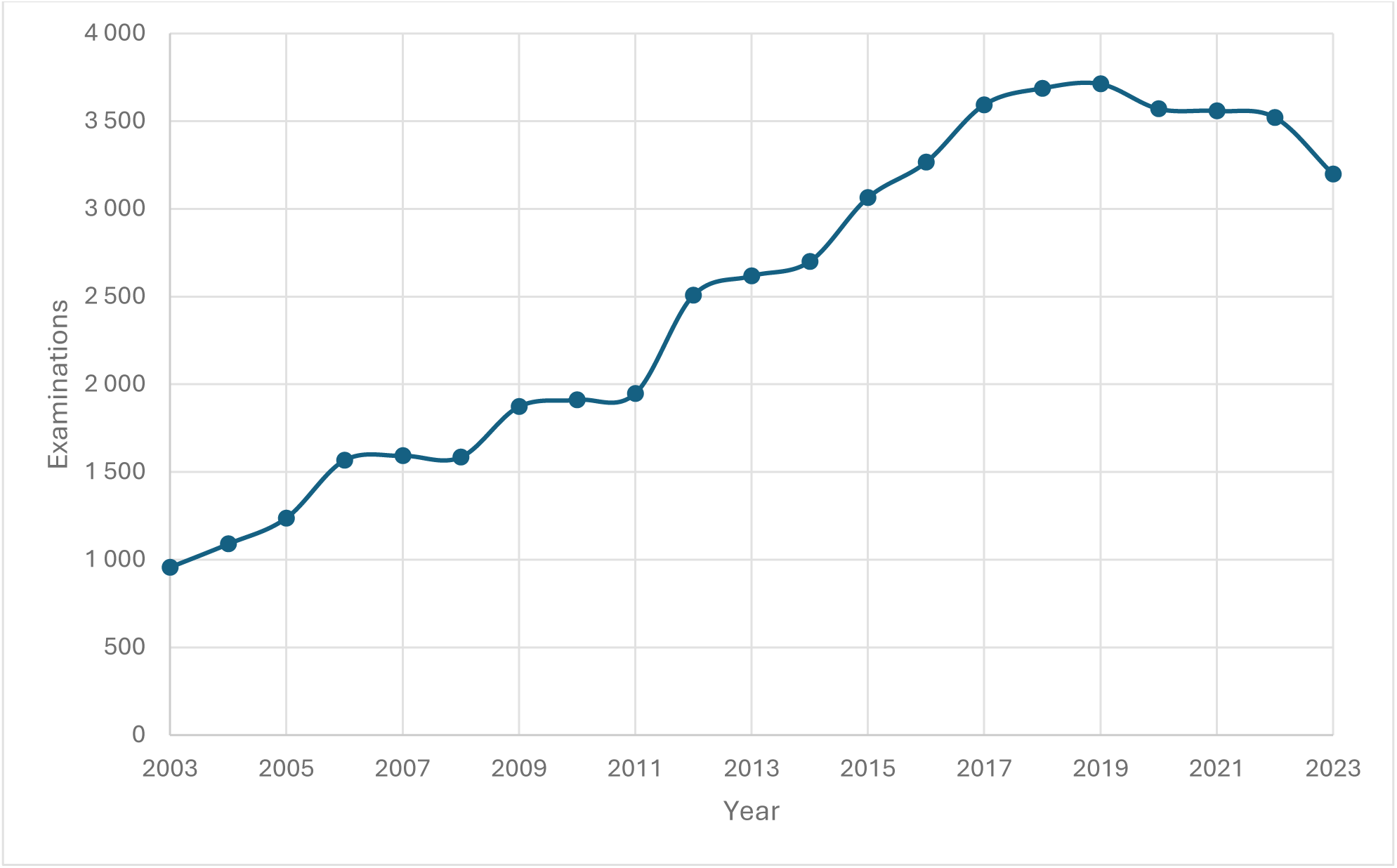
Number of head MRI examinations for patients over 50 years by year, obtained from the PACS of the Wellbeing services county of North Savo, Kuopio, a large regional public healthcare provider in Eastern Finland. PACS started to serve Kuopio University Hospital in 2003, and district hospitals and primary care units joined the regional PACS between 2006-2011.

**Supplementary Figure 2:**
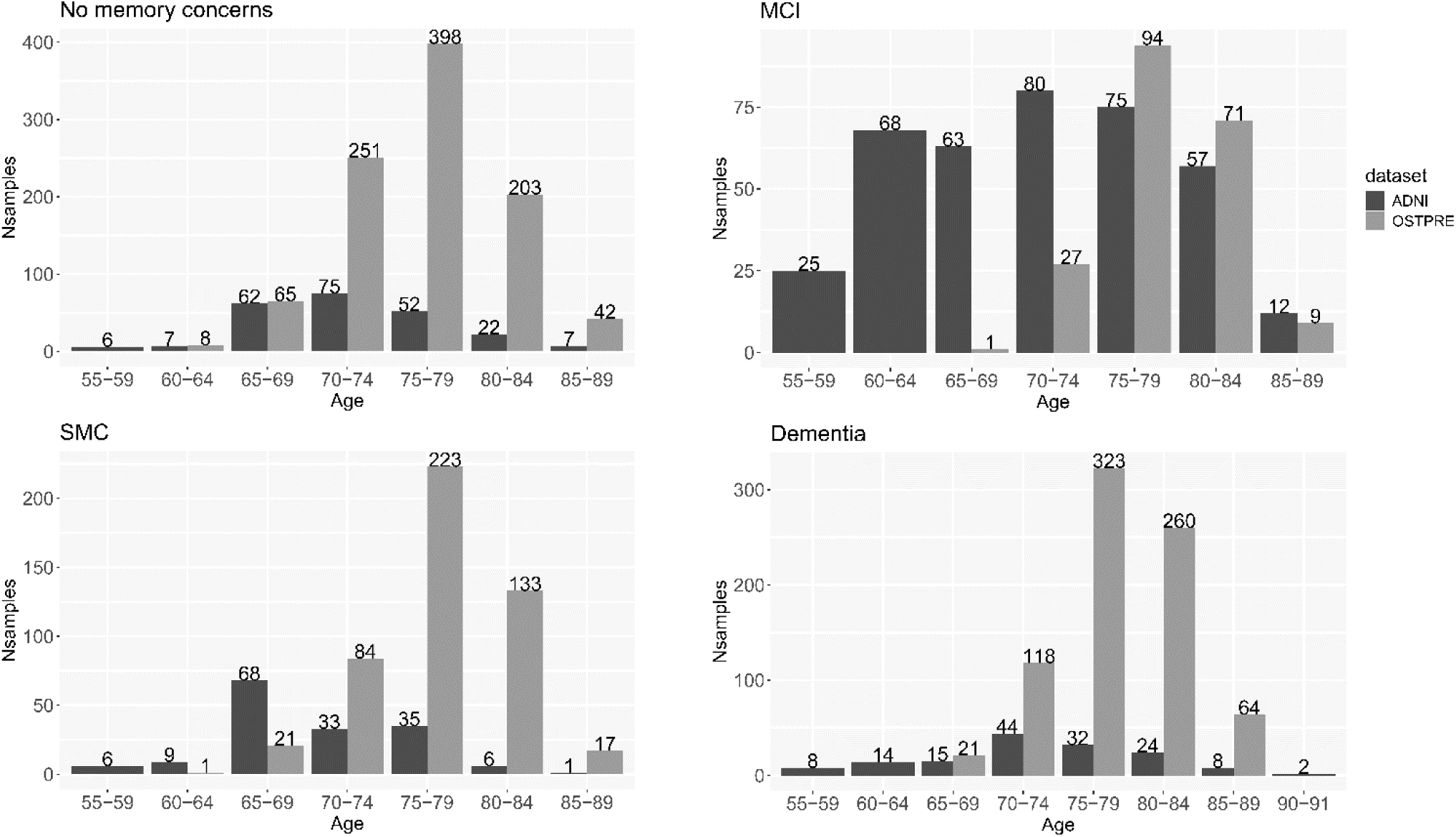
Age distribution across diagnostic groups in ADNI and OSTPRE datasets

**Supplementary Figure 3.**
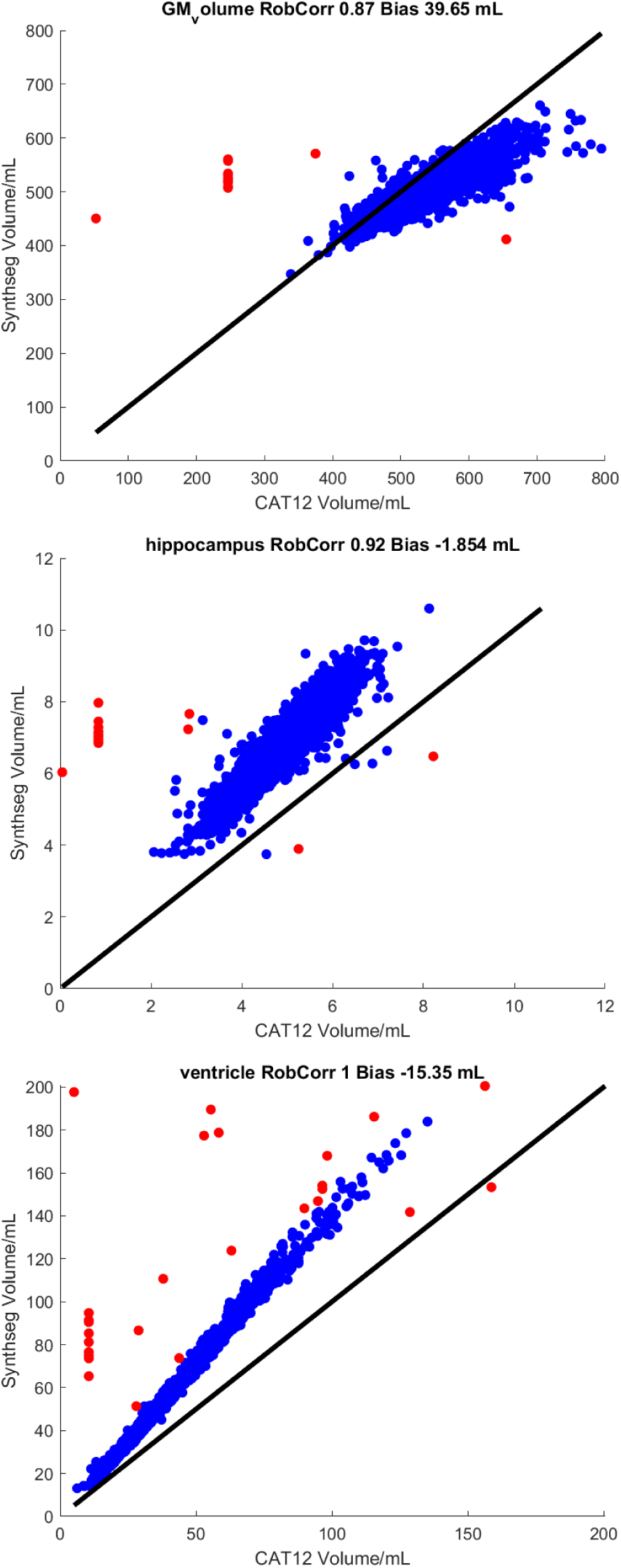
CAT12 regional volumes compared to SynthSeg 2.0 volumes with outliers indicated in red. We subjected the scans and segmentations corresponding outlying volumes to further manual quality check.

**Supplementary Table 1:** Number of head MRI examinations for patients over 50 years for each MRI scanner models between years 2003-2023, obtained from the PACS of the Wellbeing services county of North Savo, Kuopio, a large regional public healthcare provider in Eastern Finland. PACS started to serve Kuopio University Hospital in 2003, and district hospitals and primary care units joined the regional PACS between 2006-2011.

| **Manufacturer** | **Model** | **Magnetic field strength** | **Scanner type** | **Number of head MRI examinations** |
| --- | --- | --- | --- | --- |
| Siemens Healthineers | MAGNETOM Avanto | 1.5T | Normal | 14823 |
| Siemens Healthineers | MAGNETOM Avanto Fit | 1.5T | Normal | 9411 |
| Philips Medical Systems | Achieva | 3T | Normal | 9389 |
| Siemens Healthineers | MAGNETOM VISION | 1.5T | Normal | 3748 |
| Siemens Healthineers | MAGNETOM Sola | 1.5T | Normal | 3259 |
| Siemens Healthineers | MAGNETOM Aera | 1.5T | Normal | 2833 |
| GE medical systems | SIGNA Artist | 1.5T | Normal | 2639 |
| GE medical systems | SIGNA HDxt | 1.5T | Mobile | 2137 |
| Siemens Healthineers | MAGNETOM Vida | 3T | Normal | 1956 |
| Siemens Healthineers | MAGNETOM ESSENZA | 1.5T | Normal | 747 |
| Siemens Healthineers | MAGNETOM VISION plus | 1.5T | Normal | 415 |
| TOSHIBA | Vantage Elan | 1.5T | Normal | 216 |
| Siemens Healthineers | MAGNETOM Sola Fit | 1.5T | Normal | 176 |
| GE medical systems | SIGNA Voyager | 1.5T | Mobile | 157 |
| GE medical systems | Optima MR360 | 1.5T | Normal | 157 |
| GE medical systems | SIGNA EXCITE | 1.5T | Mobile | 140 |
| Other or unknown | Scans imported to PACS from other hospitals | Typically 1.5T | Typically normal | 577 |

**Supplementary Table 2.** Twenty the most common units ordering head MRI examinations for 50+ patients and number of examinations 2003-2023, obtained from the PACS of the Wellbeing services county of North Savo, Kuopio, a large regional public healthcare provider in Eastern Finland. PACS started to serve Kuopio University Hospital in 2003, and district hospitals and primary care units joined the regional PACS between 2006-2011.

| **Department/Unit** | **Number of**<br>**Examinations** |
| --- | --- |
| Neurology outpatient clinic | 15023 |
| Neurology inpatient ward | 4730 |
| Neurosurgery inpatient ward | 4467 |
| Neurosurgery outpatient clinic | 4438 |
| ED clinic | 4424 |
| Ophthalmology outpatient clinic | 2757 |
| Ear-nose-throat outpatient clinic | 2321 |
| Primary care health centre | 2025 |
| Geriatric outpatient clinic | 1202 |
| Hearing and balance centre | 1141 |
| Intensive care inpatient ward | 1066 |
| ED ward | 1065 |
| Oncology outpatient clinic | 851 |
| Endocrinology outpatient clinic | 842 |
| Heart outpatient clinic | 564 |
| Epilepsy centre | 485 |
| Oncology inpatient ward | 416 |
| University research scans | 411 |
| Internal medicine ward | 309 |
| Other | 3645 |

**Supplementary Table 3.**
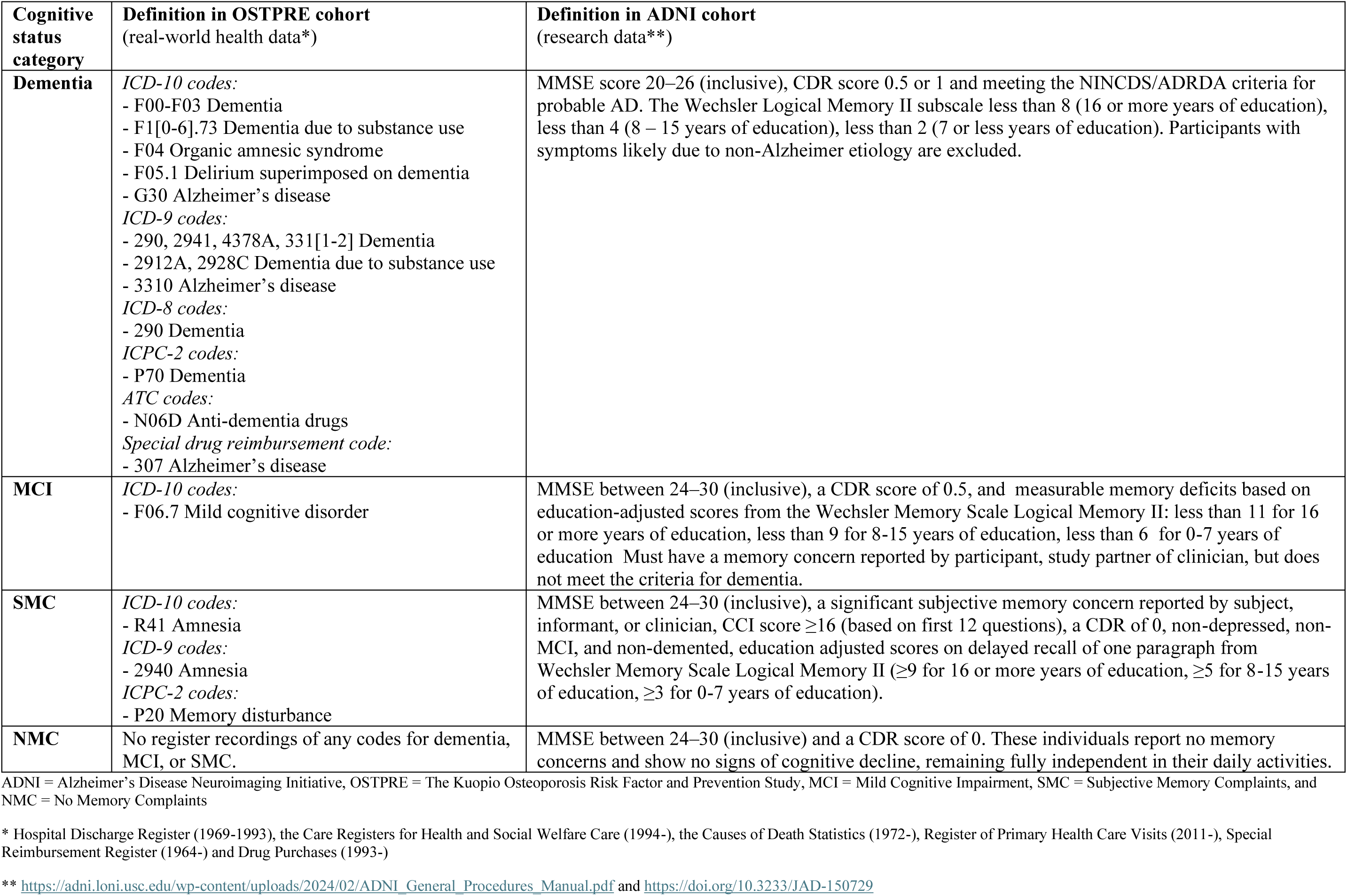
Description of cognitive status categories in the OSTPRE and ADNI cohorts.

| **Cognitive status category** | **Definition in OSTPRE cohort**<br>**(real-world health data\*)** | **Definition in ADNI cohort**<br>**(research data\*\*)** |
| --- | --- | --- |
| **Dementia** | *ICD-10 codes:*<br>- F00-F03 Dementia<br>- F1[0-6].73 Dementia due to substance use<br>- F04 Organic amnesic syndrome<br>- F05.1 Delirium superimposed on dementia<br>- G30 Alzheimer's disease<br>*ICD-9 codes:*<br>- 290, 2941, 4378A, 331[1-2] Dementia<br>- 2912A, 2928C Dementia due to substance use<br>- 3310 Alzheimer's disease<br>*ICD-8 codes:*<br>- 290 Dementia<br>*ICPC-2 codes:*<br>- P70 Dementia<br>*ATC codes:*<br>- N06D Anti-dementia drugs<br>*Special drug reimbursement code:*<br>- 307 Alzheimer's disease | MMSE score 20–26 (inclusive), CDR score 0.5 or 1 and meeting the NINCDS/ADRDA criteria for probable AD. The Wechsler Logical Memory II subscale less than 8 (16 or more years of education), less than 4 (8 – 15 years of education), less than 2 (7 or less years of education). Participants with symptoms likely due to non-Alzheimer etiology are excluded. |
| **MCI** | *ICD-10 codes:*<br>- F06.7 Mild cognitive disorder | MMSE between 24–30 (inclusive), a CDR score of 0.5, and measurable memory deficits based on education-adjusted scores from the Wechsler Memory Scale Logical Memory II: less than 11 for 16 or more years of education, less than 9 for 8-15 years of education, less than 6 for 0-7 years of education. Must have a memory concern reported by participant, study partner or clinician, but does not meet the criteria for dementia. |
| **SMC** | *ICD-10 codes:*<br>- R41 Amnesia<br>*ICD-9 codes:*<br>- 2940 Amnesia<br>*ICPC-2 codes:*<br>- P20 Memory disturbance | MMSE between 24–30 (inclusive), a significant subjective memory concern reported by subject, informant, or clinician, CCI score $\geq 16$ (based on first 12 questions), a CDR of 0, non-depressed, non-MCI, and non-demented, education adjusted scores on delayed recall of one paragraph from Wechsler Memory Scale Logical Memory II ($\geq 9$ for 16 or more years of education, $\geq 5$ for 8-15 years of education, $\geq 3$ for 0-7 years of education). |
| **NMC** | No register recordings of any codes for dementia, MCI, or SMC. | MMSE between 24–30 (inclusive) and a CDR score of 0. These individuals report no memory concerns and show no signs of cognitive decline, remaining fully independent in their daily activities. |

ADNI = Alzheimer's Disease Neuroimaging Initiative, OSTPRE = The Kuopio Osteoporosis Risk Factor and Prevention Study, MCI = Mild Cognitive Impairment, SMC = Subjective Memory Complaints, and NMC = No Memory Complaints
\* Hospital Discharge Register (1969-1993), the Care Registers for Health and Social Welfare Care (1994-), the Causes of Death Statistics (1972-), Register of Primary Health Care Visits (2011-), Special Reimbursement Register (1964-) and Drug Purchases (1993-)
\*\* https://adni.loni.usc.edu/wp-content/uploads/2024/02/ADNI_General_Procedures_Manual.pdf and https://doi.org/10.3233/JAD-150729

